# Histopathology and proteomics are synergistic for High-Grade Serous Ovarian Cancer platinum response prediction

**DOI:** 10.1101/2024.06.01.24308293

**Authors:** Oz Kilim, Alex Olar, András Biricz, Lilla Madaras, Péter Pollner, Zoltán Szállási, Zsofia Sztupinszki, István Csabai

## Abstract

Patients with High-Grade Serous Ovarian Cancer (HGSOC) exhibit varied responses to treatment, with 20-30% showing *de novo* resistance to platinum-based chemotherapy. While hematoxylin-eosin (H&E) pathological slides are used for routine diagnosis of cancer type, they may also contain diagnostically useful information about treatment response. Our study demonstrates that combining H&E-stained Whole Slide Images (WSIs) with proteomic signatures using a multimodal deep learning framework significantly improves the prediction of platinum response in both discovery and validation cohorts. This method outperforms the Homologous Recombination Deficiency (HRD) score in predicting platinum response and overall patient survival. The study sets new performance benchmarks and explores the intersection of histology and proteomics, highlighting phenotypes related to treatment response pathways, including homologous recombination, DNA damage response, nucleotide synthesis, apoptosis, and ER stress. This integrative approach has the potential to improve personalized treatment and provide insights into the therapeutic vulnerabilities of HGSOC.

## 0 Introduction

20%–30% of HGSOC patients have treatment-refractory disease at diagnosis and have a poor prognosis ^1^. Patients with refractory disease experience the toxicity of platinum-based chemotherapy without benefit. Due to their rapid disease progression, such cases are commonly excluded from participating in clinical trials. Lack of mechanistic understanding significantly impedes developing treatment strategies to overcome first line treatment resistance. Conversely, improved prediction models for treatment resistance could significantly enhance clinical decision-making, allowing for more effective treatment approaches that minimize exposure to potentially non-effective therapies and side effects, while ensuring that patients who are likely to respond to a treatment are identified.

Homologous recombination-deficiency (HRD) plays a major role in response to platinum-based treatments and PARP-inhibitors. Cases with HRD can be identified using mutational-signature based signatures utilizing next generation sequencing-based methods including the HRD-score ^2^, the FDA-approved methods (Myriad myChoice CDx^3^ and FoundationFocus CDxBRCA LOH^4^) and WGS-based HRDetect ^5^ or WES-retrained HRDetect ^6^ and functional assays. (e.g. RAD51 foci). Although recent publications investigated the possibility of predicting HRD-status using whole slide images ^7,8^ and predicting response to platinum-based treatments using WSI or proteomics data ^9–11^, their predictive power has not reached the level of clinical utility.

Investigating the proteome is informative of how tumors respond to platinum-based treatments. Recent studies in proteogenomics have revealed that certain patterns of protein expression and modifications are linked to patient survival rates and outcomes in cases of High-Grade Serous Ovarian Carcinoma (HGSOC) ^10,12,13^. Additionally, Chowdhury et.al^9^ described a signature of 64 proteins that can predict with high specificity which patients might develop resistance to initial platinum therapy. However, this approach captures only part of the complex interaction between cancer cells and treatment. The spatial organization of these cells within the tumor microenvironment, a key aspect that proteomic analyses often overlook, also significantly influences treatment response. Understanding the spatial distribution of proteins and their interaction with the tumor environment adds another layer of insight, potentially identifying mechanisms that drive therapy resistance and efficacy.

Recent advances in computational pathology demonstrate great potential for treatment response prediction using weakly supervised deep learning models. These computer vision algorithms can use H&E-stained Whole Slide Images (WSIs) to predict the outcomes of treatments like bevacizumab in ovarian cancer ^14^. Additionally, these models can determine Homologous Recombination Deficiency (HRD) status ^7,8,15^. This capacity to derive significant clinical insights directly from WSIs suggests that when combined with proteomic data, which offers a detailed molecular landscape of tumor behavior, we can significantly improve predictive models.

Since WSI, proteomic signatures or DNA repair deficiency status are all predictive of various aspects of ovarian cancer biology, it is possible that their “fusion” may produce even superior predictive performance.

Early and late fusion models are two approaches in multimodal data fusion and analysis. Early fusion is similar to a multidisciplinary consultation where an oncologist, radiologist, and genetic counselor collaborate from the start, pooling their expertise to quickly form a comprehensive view of a patient’s health. This approach is useful when the interplay among various medical data types is essential for accurate diagnosis and treatment planning. In contrast, late fusion is akin to sequential specialist consultations. Each expert examines the patient independently, contributing insights from their respective fields in detail and later integrate these findings. This method allows for in-depth analysis of each data type, ensuring that unique insights are captured before they are synthesized into a diagnosis or treatment plan.

Our study evaluates early and late fusion models by benchmarking recent methodological advances in multi-modal integration of histopathology and omics data ^16–20^. We compare these models across metrics of performance and interpretability. Furthermore, multi-modal fusion provides opportunities for automatically identifying biomarkers related to treatment resistance. The way these models combine different modalities is designed to be inherently interpretable. Therefore, the integration of biological knowledge into the models facilitates detailed investigations at the patient level into the significance of specific histopathology morphologies, pathways and proteins.

Our study is the first to integrate proteomics and histopathological images and show that these modalities are synergistic for platinum response treatment. The complementarity between the global, spatial context provided by WSIs and the mechanistic information. Furthermore, we show that these models can be probed to provide fine-grained interpretability to better understand tumor response at a mechanistic level.

## 1 Results

### 1.1 Combination of WSI with proteomics significantly increases the accuracy of response prediction

Both the Cancer Genome Atlas Program (TCGA) ^12^ and Proteogenomic Translational Research Centers High-Grade Serous Ovarian Carcinoma (PTRC-HGSOC) ^9^ datasets included patients with paired H&E WSI and proteomics data, along with documented responses to platinum chemotherapy. This enabled an investigation into the effectiveness of multi-modal histology-proteomics deep learning compared to their unimodal counterparts for this task.

We first trained a clustering-constrained attention multiple instance learning (CLAM) ^21^ model to predict response to platinum therapy in ovarian cancer based on WSI data alone. The models were trained on one of the data sets available (TCGA or PTRC-HGSOC, see methods for description of data sets) and validated on the other data set. As Table. 1 shows, the predictive power of this method using only pathological images was rather limited independent of whether primary or metastatic tumors were considered. Similarly, we trained a classical machine learning ensemble model based on the proteomics “Chowdhury Signature” ^9^ (methods 3.6.1) on one of the two data sets (TCGA-OV or PTRC-HGSOC) and evaluated the predictive power on the other data set (full split descriptions in supplementary figure 3.a. The proteomics-based predictor performed better than the WSI-based predictor (Table. 1).

**Figure 1.**
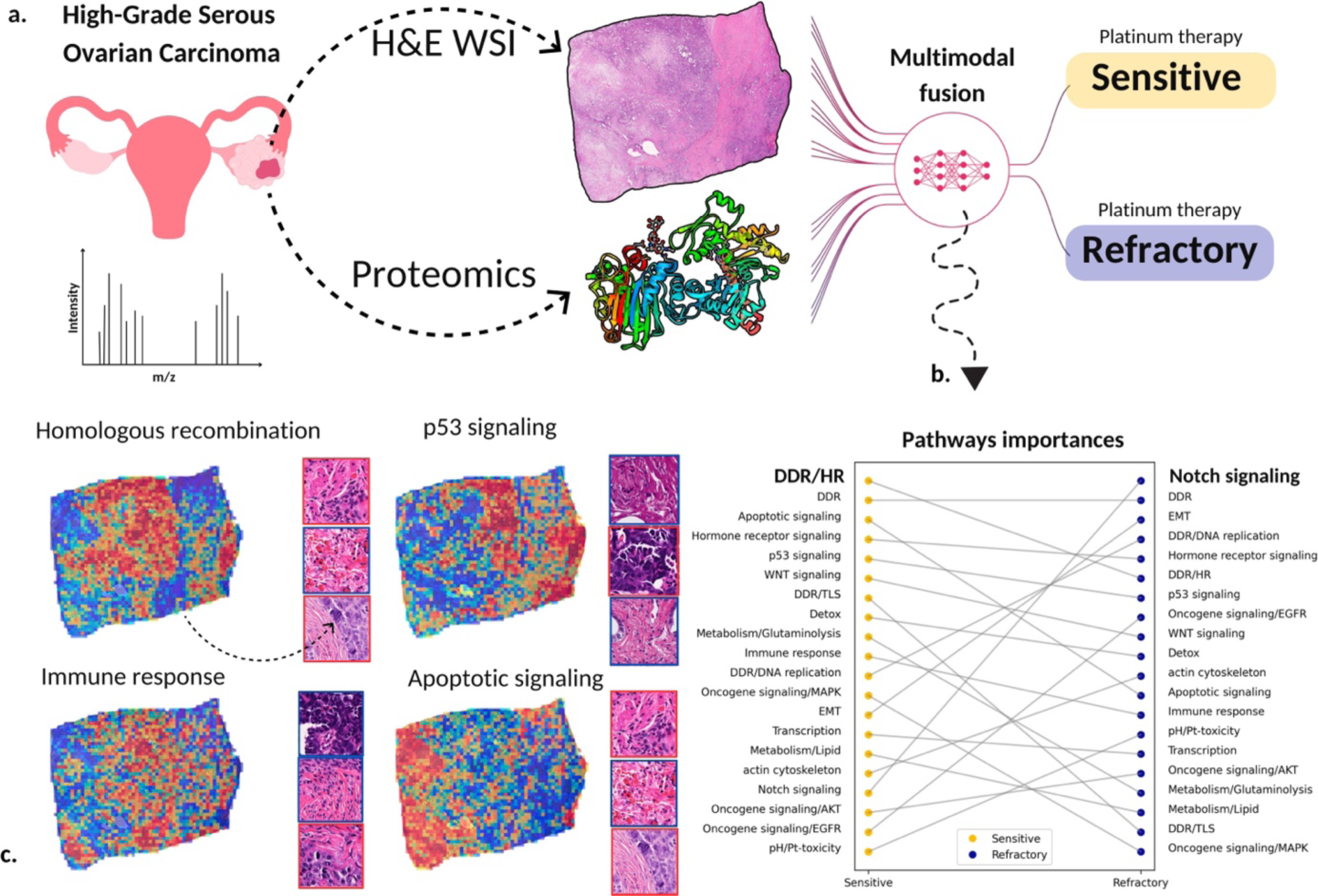
Experiment overview. **a**. Training various multi-modal deep learning models on paired H&E whole slide images and proteomics measurements to predict High-Grade Serous Ovarian Cancer tumor sensitivity to platinum-based therapy. **b**. Local and global interpretability of learned features. Ranking of importance of pathways in platinum response. **c**. Visualization of learned co-attention between bulk proteomics and whole slide image patches.

**Table 1.** The table displays separated reported Area Under the Curve (AUC) scores for primary and metastatic tumors with the multimodal PorpoiseMMF ^18^ model + Chowdhury signature (methods 3.6.1). The histopathology model is the CLAM ^21^ architecture, and the proteomics model is from the “Chowdhury signature” trained with an ensemble model as described in Chowdhury et.al ^9^. Multimodal models outperform their unimodal counterparts. Test AUCs represent the mean AUC obtained from the ensembled predictions of the five models. Error estimates are obtained through bootstrapping with 1,000 samples, providing an assessment of variability. For validation scores, we report the average AUCs across the folds, with the standard deviation of these AUCs representing the reported errors.

|  | Model | HGSOC Train |  | TCGA Train |  | HGSOC Train |  |
| --- | --- | --- | --- | --- | --- | --- | --- |
|  |  | TCGA Test | 5 fold CV | HGSOC Test | 5 fold CV | UAB Test | 5 fold CV |
| Primary | Histopathology | 0.566±0.052 | 0.655±0.159 | 0.528±0.051 | <u>0.586±0.127</u> | 0.523±0.105 | 0.567±0.099 |
|  | Proteomics | <u>0.61±0.05</u> | <u>0.696±0.115</u> | <u>0.755±0.041</u> | 0.552±0.085 | <u>0.558±0.094</u> | <u>0.723±0.129</u> |
|  | **Multimodal** | **0.752±0.044** | **0.796±0.105** | **0.835±0.039** | **0.613±0.136** | **0.84±0.071** | **0.781±0.14** |
| Metastatic | Histopathology | 0.486±0.056 | 0.68±0.138 | 0.431±0.046 | <u>0.554±0.122</u> | <u>0.769±0.08</u> | 0.648±0.172 |
|  | Proteomics | <u>0.54±0.053</u> | **0.873±0.058** | <u>0.566±0.046</u> | 0.542±0.08 | 0.665±0.092 | **0.75±0.15** |
|  | **Multimodal** | **0.704±0.048** | <u>0.79±0.076</u> | **0.698±0.042** | **0.608±0.129** | **0.798±0.063** | <u>0.727±0.21</u> |

Next, we asked whether we could derive a more accurate predictor if we combine patient-paired H&E WSIs and proteomics data using multi-modal deep learning frameworks ^18–20^. The model combining WSI and proteomic data results in a significant increase in model performance. (Table. 1). Multi-modal benchmarks consistently outperformed both WSI-only and proteomics-only models predicting treatment resistance to platinum chemotherapy. This finding was robust across various training and testing setups (See methods section 3.1 and supplementary Figure. S3.a). This suggests that there exist multi-modal features invariant across cohorts that correlate well with tumor sensitivity to treatment. The test AUC results pertain solely to cohorts from held-out sites, ensuring the mitigation of potential biases and validating the generalization of discovered features ^22^. Table. 1 presents the best performing PorpoiseMMF ^18^ model (supplementary section 3.5) performance for the multimodal model. Tables containing the results for all benchmarked models can be found within the supplementary material (supplementary Tables S1-S4).

The results for primary samples shown in Table 1 demonstrate that overall, proteomics-only models slightly outperform WSI-only models. When training on PTRC-HGSOC ^9^ primary tumor samples and testing on TCGA samples, the multi-modal model achieves an AUC (Area Under the Curve) of 0.752. This is a significant improvement over the proteomics-only model, which has an AUC of 0.61. This represents a 14% increase (t=6.24, p=0.00336). When training on TCGA tumor samples and testing on PTRC-HGSOC primary tumor samples, the multi-modal model achieves an AUC of 0.835. This represents a 13.6% increase over the proteomics-only model, which has an AUC of 0.755 (t=4.14, p=0.0144). Finally, when training on the FHCRC+Mayo sub-cohorts (figure. 2.) of PTRC-HGSOC primary tumor samples and testing on UBC primary tumor samples, the multi-modal model achieves an AUC of 0.84. This represents a 19.9% increase over the proteomics-only model, which has an AUC of 0.558 (t=7.0, p=0.00219).

**Figure 2.**
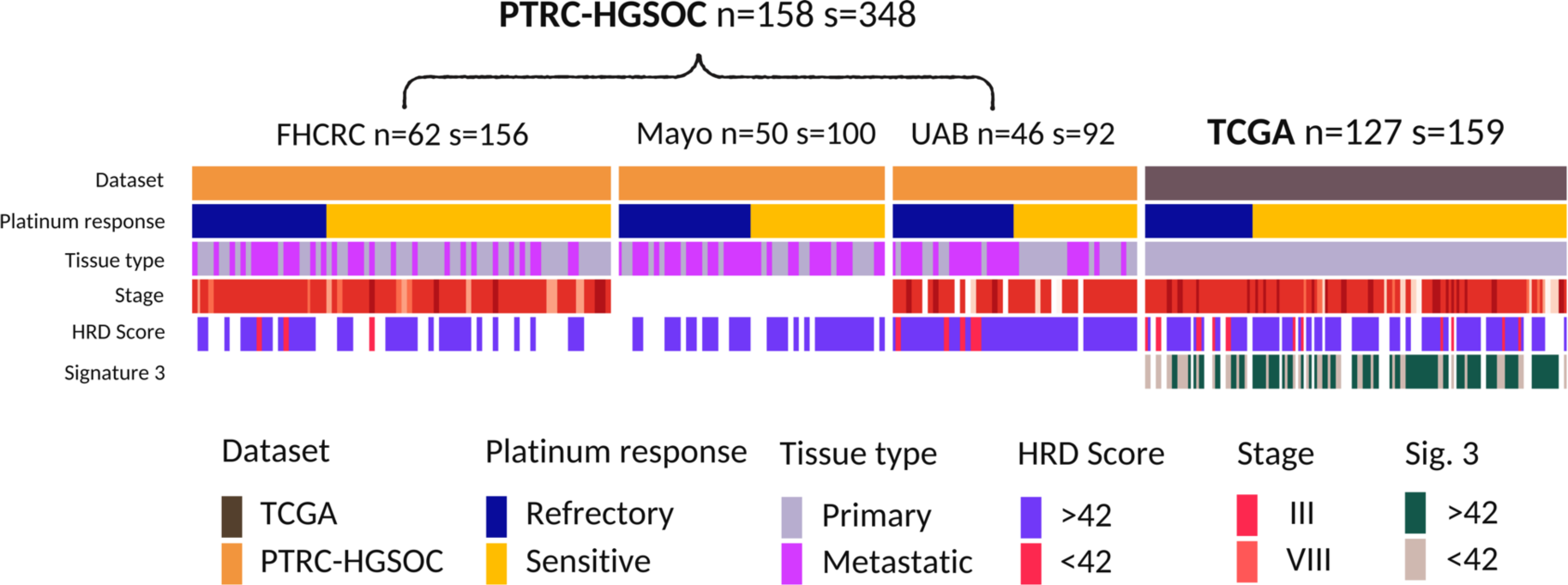
Data overview for the study. Three hospitals constitute the “discovery” cohort, named Proteogenomic Translational Research Centers High-Grade Serous Ovarian Carcinoma (PTRC-HGSOC). Proteomics from the CPTAC-OV study was used for the independent TCGA cohort.

Focusing on metastatic cases separately, Table 1 demonstrates that overall, proteomics-only models slightly outperform WSI-only models, and all test results are slightly lower than primary trained models, as TCGA samples are only from primary tumor sites. When training on PTRC-HGSOC metastatic tumor samples and testing on TCGA samples, the multi-modal model achieves an AUC of 0.704, representing an 8.2% increase over the proteomics-only model, which has an AUC of 0.54 (t=2.63, p=0.058). Training on TCGA tumor samples and testing on PTRC-HGSOC metastatic tumor samples, the multi-modal model achieves an AUC of 0.698, representing a 13.7% increase over the proteomics-only model, which has an AUC of 0.566 (t=2.58, p=0.0614). Finally, when training on the FHCRC+Mayo sub-cohorts of PTRC-HGSOC metastatic tumor samples and testing on UBC metastatic tumor samples, the multi-modal model achieves an AUC of 0.798, representing a 16.5% increase over the proteomics-only model, which has an AUC of 0.665 (t=3.33, p=0.029).

When modeling histopathology-proteomics data, the experimental setup involves selecting the model architecture, the proteomic input structure (methods 3.6), and the histopathology patch embeddings (methods 3.4). We evaluated how changes in each of these choices impacted the performance in predicting platinum response.

Comparing multimodal model architectures, we found that PorpoiseMMF ^19^ outperformed MCAT ^20^ and SurvPath ^19^ in overall model performance. The former being a late fusion model and the latter two being early fusion models. In terms of proteomic groupings, Chowdhury signature (methods 3.6.1) > CPTAC signature (methods 3.6.2) > RM pathways (methods 3.6.4) > PRG pathways (3.6.3). Comparing pretrained histopathology image patch embedding networks, our SSL trained ovarian cancer-specific DINO-OV (supplementary Table S5 for pre-training data and supplementary section 3.8 for SSL details), UNI^23^ and CTransPath ^24^ performed similarly across the tasks (supplementary Tables S1-S4).

Although primary and metastatic cancers show many similarities, they demonstrate large differences in response and prognosis. We found that training the models separately for the primary and metastatic cohorts yields better performance. Based on these findings we also recommend this practice for similar studies.

### 1.2 WSI+proteomics compared to genetics-based models

Loss of certain biological functions, such as the capacity of homologous recombination-based DNA repair, has been known to be associated with response to DNA damaging therapy in ovarian cancer ^25^. We compared our WSI+proteomics-based model predictions against the predictive power of genomics-based methods of detecting homologous recombination deficiency status. The FDA-approved Myriad HRD-score assesses genomic alterations to indicate the presence of HRD. The FoundationOne CDx test offers comprehensive genomic profiling, including TMB and genome-wide LOH (loss of heterozygosity) status, which correlates with HRD status. Research tools like HRDetect ^26^ utilize whole-genome sequencing and lasso regression to predict HRD with high sensitivity. Finally, SigMA exposes notable mutational signatures associated with HRD where Signature 3 ^27^ is of special interest.

For our studied cohorts, the HRD-score, Signature 3 and HRDetect scores were available for n=98 TCGA cases. The WSI+proteomics model outperformed the HRD-score, HRDetect, and Signature 3 for the TCGA cohort in predicting tumor response to platinum-based therapy. We found that a linear combination of our WSI+proteomics model and HRD-score showed a significant increase in performance over the pure HRD-score model (DeLong test Z-statistic: −3.113, P-value: 0.002) (See Figure 3.a, Figure 3.b, and supplementary Figure 4.c). Furthermore, our model predictions were uncorrelated with the HRD-score predictions implying genomics-based and histo-proteomics information is complementary containing little mutual information regarding therapy response prediction (See Figure 3.c). Performing a more granular investigation into model predictions over the cohorts, we visualize AUC scores for the cohorts distinguished by various clinical variables (See Figure 3.d)

**Figure 3.**
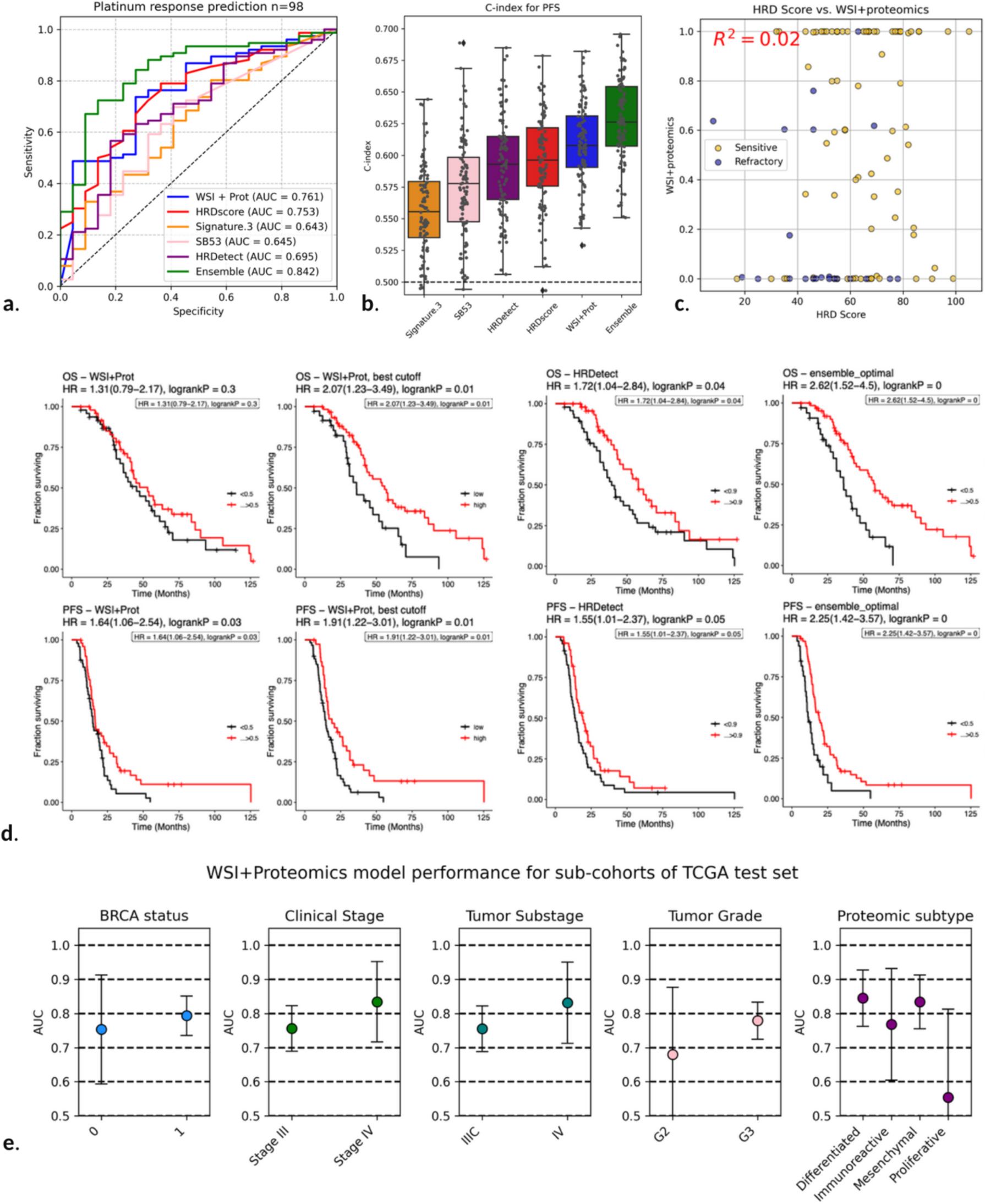
Model performance compared to WES-based methods. **a**. ROC-AUC curve for patient aggregated scores in response prediction. Our WSI+proteomics model outperforms the FDA-approved Myriad HRD-score (LOH + TAI + LST) ^34^ and can be ensembled with this model for a significant improvement over both. **b**. C-Index for our models compared to geneomics models. **c**. HRD-score does not correlate with WSI+protemics predictions highlighting the independence of signal between WES vs Histopathological and Proteomic modalities in predicting platinum response. **d**. Survival curves for TCGA cohort. Our WSI+proteomics model significantly outperforms the use of bonified BRCA status, HRDetect in separating risk groups and predicting PFS and PFI of patients. **e**. WSI+proteomics model scores for clinical subgroupings of the TCGA-OV test set.

#### 1.2.1 Predicting survival – Prognostic evaluation

To further evaluate and validate the utility of our multi-modal WSI+proteomics model we conducted survival analysis, comparing the prognostic and predictive potential of the model to other, previously established measurements. In case of the TCGA-OV cohort Cox proportional hazard models were used to determine hazard ratios and compare survival (Overall Survival (OS), DSS, PFI, PFS) and HRD-score, HRDetect, WSI-only, proteomics-only, WSI+proteomics and ensembled models using median cutoff or best cutoff, and the plots for and Kaplan-Meier analysis was performed with the survival R package and Overall Survival (OS), Disease-Specific Survival (DSS), Progression-Free Interval (PFI), Progression-Free Survival (PFS), and Cox proportional hazards models were utilized to determine Hazard Ratios for the validation and TCGA-OV cohort (See Figure 3.d). Our WSI+proteomics multi-modal model outperformed HRDetect for survival prediction its ability to separate risk groups. For OS, logrank HRDetect p=0.04 WSI+proteomics p=0.01. For PFS logrank HRDetect p=0.05 WSI+proteomics p=0.01. This is further shown by comparing C-Index values for PFS prediction across the models (Figure 3.b). Our models were trained to predict the response to treatment rather than individual survival or risk. Nonetheless, as the model demonstrates prognostic efficacy, it further validates its capacity to distinguish between patients with favorable and unfavorable responses accurately. The full analysis is shown in Supplementary Figure. S2.

### 1.3 Neural network attention maps reveal interpretable spatial distributions from bulk proteomics

As shown for the WSI in Figure. 4.a, the uni-modal WSI-only CLAM ^21^ allows us to visualize patch importances, or “attentions,” on the WSI, creating an “attention heatmap” (Figure. 4.b). This visualization provides insight into the model’s decision-making process. Patches with high attention are crucial for classifying the entire slide. For instance, if tumor areas of the WSI are highlighted on the attention map for a task that relies on known tumor localized histopathological features, it may validate that the model aligns with pathologists’ thinking. Additionally, when we lack documented histopathological features for a task, these heatmaps can help us discover important features the model considers significant. This is particularly relevant for predicting platinum response, where histopathology is not a primary biomarker for decision-making.

**Figure 4.**
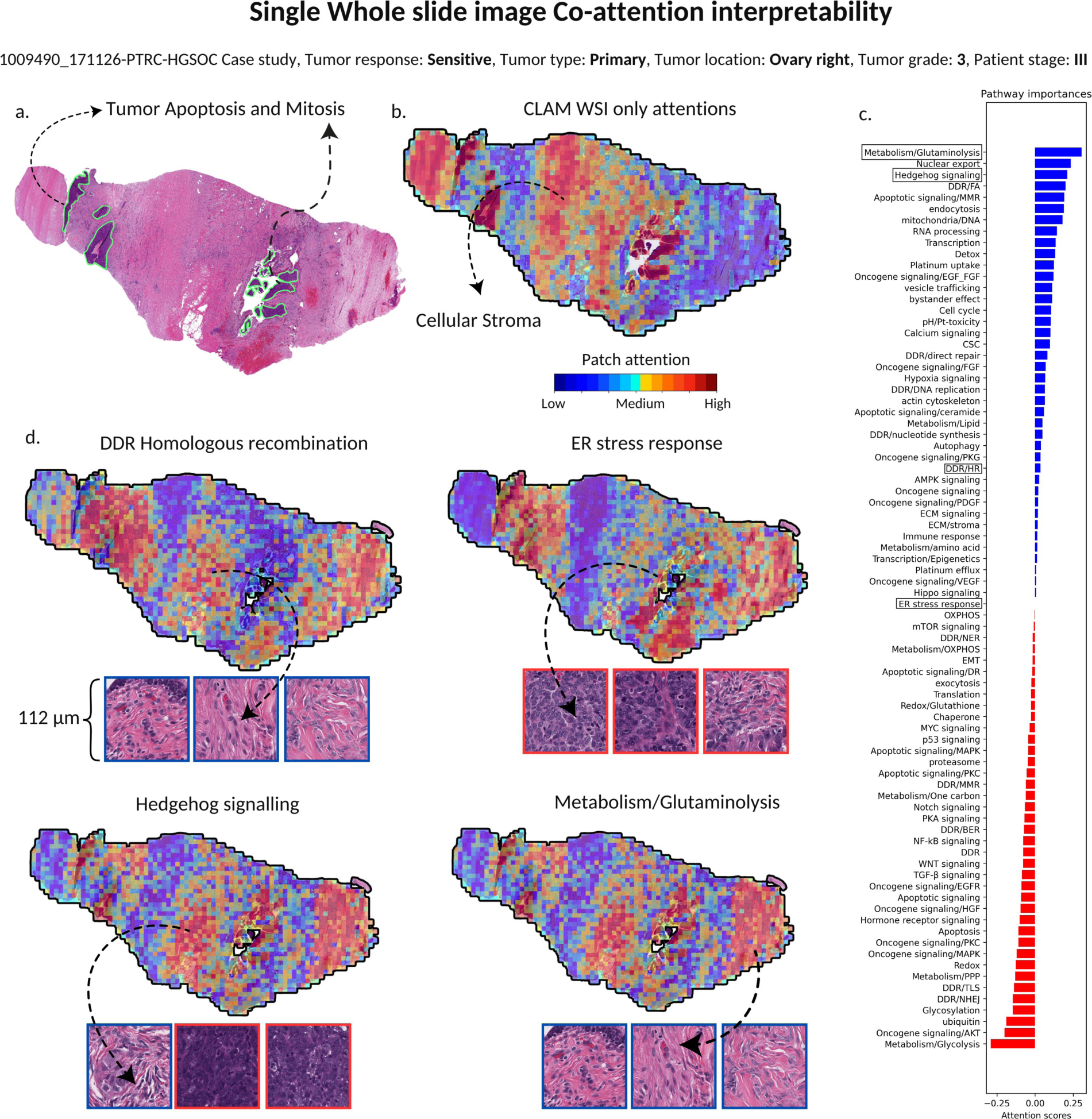
Model interpretability example for a WSI+proteomics pair. **a.** WSI with tumor areas segmented. **b.** The MCAT model assigns tumor histopathology patches high attention as well as some specific non-tumor or adjacent cellular stroma. **c.** Model ranking of pathway importance in the decision-making process for the model for this WSI Proteomics paired data. **d.** 4 out of the 80 MCAT model co-attention maps where the model projects the non-spatial proteomics data into the spatial domain on the WSI.

Extending this concept into multimodal models, MCAT ^20^ and SurvPath ^19^ enable the visualization of more complex “co-attention heatmaps.” For each biological pathway, the model generates one “gene-guided visual concept” ^20^for each patch in the WSI. These highlight important visual features integrated with proteomic data. The fact that the “co-attention heatmaps” in Figure 4.d can reveal spatial features may seem to be surprising at first sight. However, this is alike deep neural network’s ability to recognize objects in images after training without detailed annotations. For an intuitive description of how images are connected to proteomic features see supplementary section 4.2.

A key strength of MCAT ^20^ and SurvPath ^19^architectures is the ability to visualize these pathway-specific attention maps over WSIs, thereby potentially discovering important morphological features related to treatment sensitivity or resistance. While the critical role of the tumor microenvironment and stroma in treatment response and disease progression is widely recognized, routine histopathological assessments often overlook these components. We found that the stroma adjacent to tumor cells frequently attracts significant attention in numerous slides and cases, underscoring the presence of valuable information within the non-tumor regions of pathological slides. In Figure. 4 we showcase an example case study. Furthermore, we validate the robustness of these generated heatmaps with an ablation study (supplementary Figure. S4.a).

### 1.4 Pathways and genes with high attention

The MCAT ^20^ and SurvPath ^19^models are pioneering efforts in integrating multimodal data to enhance model interpretability within the domain of clinical outcomes. These models employ an early fusion technique that incorporates pathways; groups of proteins with interconnected functions, as fundamental components of their architecture. This approach leverages learnable embeddings to represent these pathways, generating a single value or “attention” that defines how specific pathways contribute to clinical endpoints. By incorporating pathway embeddings, these models facilitate a method of ranking and visualizing the relative importance of different pathways in relation to a clinical outcome. This visualization is achieved through the analysis of pathway embeddings’ weights, offering insights into which pathways are most critical for the model’s predictions (Figure 4.c).

Furthermore, the models assign “feature importances” to individual protein expression values within each pathway group. These importances are derived from integrated gradient values ^28^, a method detailed in the supplementary section. Using both interpretability measures gives a two-tiered overview or “hierarchical explainability,” where both the pathway level and the protein level within pathways are illuminated in terms of their contribution to the model’s decision-making process.

To assess whole cohort trends in pathway and proteomic significance, we aggregate these feature importances across the PTRC-HGSOC cohort. This analysis is conducted separately for tumors classified as sensitive and refractory, providing a broad overview of unique proteomic and pathway dynamics across these two categories. The results, highlight the prominence of the HRD pathway in sensitive tumors. In refractory tumors, pathways related to DDR nucleotide synthesis, apoptosis, ER stress, and P53 signaling are identified as significantly influential.

To further investigate the relative importance of individual domain-specific pathways related to HGSOC platinum response we found that the BIOCARTA ATR-BRCA ^29^ pathway was the most important in both sensitive and refractory cohorts (Figure. 5.a, 5.b). Non-homologous end joining was ranked second for refractory samples and Base excision repair was second highest for the sensitive cohort. Within the BIOCARTA ATR-BRCA ^29^ pathway, we found that ATR and CHEK1 had the largest influence on model decision-making for sensitive tumors. For the refractory cohort, the models were most sensitive to CHEK1 and TP53 expression values. See Figure. 5 for the top 5 pathways and top proteins for each tumor response cohort and supplementary Figure. S1 for the ranking of all pathways and proteins.

**Figure 5.**
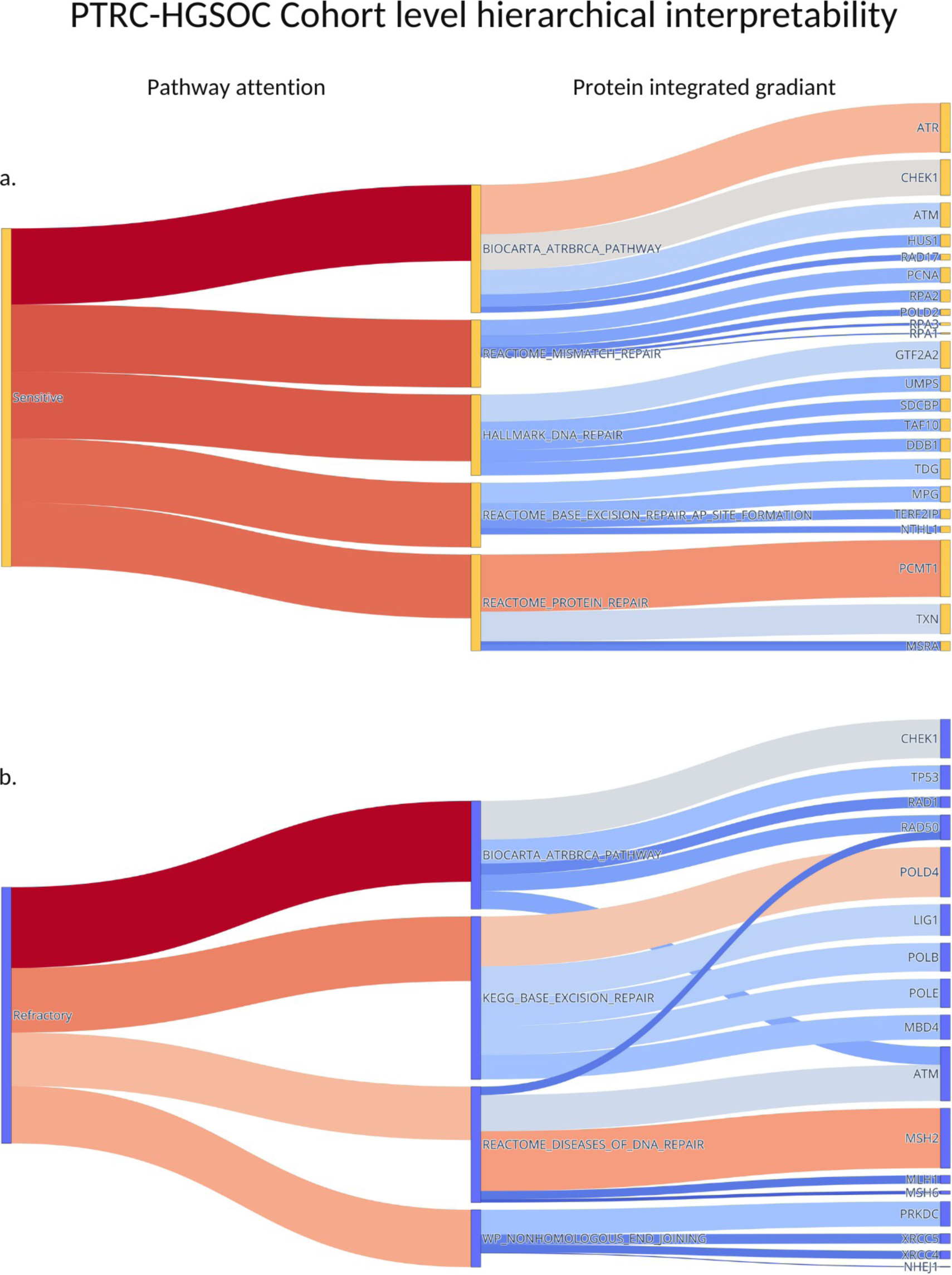
Cohort aggregated global interpretability visualized from the MCAT model. Sankey diagrams where weights connecting cohorts to the top 5 most important pathways are the mean model weights. Connections between pathways and proteins represent the four proteins in each given pathway with the largest integrated gradients. Wider connecting lines represent higher impertinences. This offers a hierarchic interpretability. See supplementary section 3.7 for the definition of integrated gradients ^28^. **a.**Sensitive pathways and proteins with high importance, **b.** Refractory pathways and proteins with high attention. See all pathways rankings in the Supplementary Figure S1.

## 2 Discussion

Debulking surgery followed by platinum-based chemotherapy has been the standard of care of HGSOC. Patients responding to first-line platinum-based therapy often have a durable remission and the clinical benefit could also be further extended by the administration of targeted agents, such as PARP inhibitors. However, platinum resistance, defined as disease recurrence within 6 months of first-line platinum-based chemotherapy, occurs in approximately 25% of cases and the prognosis of such cases is worse with a median survival of 9–12 months ^30,31^. Such cases may benefit from novel therapy such as the recently approved antibody drug conjugate Mirvetuximab Soravtansine^32^. Accurate prediction of resistance to platinum-based therapy may allow the early administration of other therapies, without waiting several months of failed platinum therapy. In principle, the surgical, debulking biopsy could be used to predict response to therapy to first-line platinum treatment, but no diagnostic method has reached the desired predictive accuracy so far.

A great number of biomarkers have been evaluated to predict platinum sensitivity in HGSOC ^33^. Some of those biomarkers were derived from genomic or transcriptomic profiling of HGSOC of known clinical outcome and others, such as quantifying HR deficiency, were developed based on biological rationale. HR deficiency can be detected by various next generation sequencing based methods such as the HRD-score ^34^, and the FDA-approved Myriad myChoice CDx ^3^ and FoundationFocus CDxBRCA LOH ^4^, which are based on similar principles. However, the predictive power of these methods was only validated for PARP inhibitor therapy ^35^, and they are not used in the decision making for first line platinum treatment.

Recently, digital pathology and associated deep learning-based computer vision algorithms emerged as an alternative diagnostic method to predict HRD status and platinum sensitivity. ^7,8,15,36^

Since molecular profiling-based, biological pathway-based and digital pathology-based predictors all showed a certain level of predictive power but none of those had sufficiently high accuracy alone, we were wondering whether these methods all capture to a certain extent different aspects of platinum response/resistance and whether those could be combined. To test this hypothesis, we decided to integrate proteomics and histopathology. We found that our models can capture non-mutual information that spans from molecular mechanisms to tissue-level manifestations of HGSOC. Proteomics provides the “what” in terms of molecular changes and potential targets, while histopathology provides the “where” and “how” these changes manifest in tissue architecture and the tumor microenvironment. Our results indicate that amalgamating information from these projections of the disease is beneficial for machine-learning models, similar to how an oncologist integrates multiple sources of information to make informed clinical decisions. Our results are concordant with other recent literature where multimodal deep learning has been shown to improve model performance in risk stratification of HGSOC using MRI, histopathology, and genomic signatures with a late fusion architecture ^37,38^. Furthermore, including clinical variables has also been shown to enhance model performance ^39^.

Our models are validated not only by various train-test configurations but also by the rediscovery of pathways known to be of high importance for HGSOC platinum response, such as Homologous Recombination Repair. Additionally, tumor regions of WSIs gather high patch attention in the WSI-only CLAM ^21^ model, validating the weakly supervised learning as a modeling decision for WSIs in this context.

We chose to focus on combining proteomic data with WSI data because there was sufficient and appropriate data available for both training and validation. Proteomics is rarely used in the diagnostic setting although reverse-phase protein arrays (RPPA) may provide a diagnostically applicable method to quantify a few hundred proteins ^40^. Combining transcriptomics or epigenomic profiling with WSI by the strategy outlined in this paper is a logical next step that may be easier to incorporate into the existing diagnostic pipelines. It is encouraging, that the combined WSI/proteomics predictor in a linear combination of HRD measures produced a predictor with an accuracy that may merit further serious consideration in the clinical setting if verified by further independent cohorts.

Variations in H&E staining routines, sample preservation, and imaging technologies across institutions pose additional challenges. We addressed these through pathology-specific self-supervised learning pre-training ^41^, effectively normalizing across such variations and ensuring our models’ applicability to diverse clinical settings. Similarly, potential discrepancies in LC-MS/MS data arising from institutional protocol differences were mitigated by comprehensive distribution overlap analyses, confirming the consistency of our biological signal capture.

## 3 Methods

### 3.1 Patient cohorts

This study aimed to predict response to platinum treatment in high-grade serous ovarian cancer by using supervised deep multi-modal learning to H&E WSIs and proteomic LC-MS/MS data. Based on literature search there were only 2 datasets with these data available: PTRC-HGSOC (158 patients 348 samples with all 3 data types) ^9^ and TCGA (127 patients 159 samples with all 3 data types)^12^. Refractory cancers were defined as those that progressed or had stable disease within 6 cycles of initial platinum/taxane therapy after initial debulking surgery^9^. Sensitive tumors were defined as those that responded to initial platinum/taxane therapy and did not progress within 2 years. All samples were treatment-naive at the time of biopsy.

### 3.2 Multimodal deep learning models

In this study we benchmarked various experimental setups. These corresponded to three histopathology patch embedding networks (methods 3.4), five model architectures (supplementary section 3.2-3.5) and 4 proteomics grouping setups (methods 3.6.1-3.6.4). Each experiment was run over 5 patient stratified folds, and three training-validation split setups. Reported test accuracies were from the ensemble of the 5 trained models’ predictions. Error estimates are obtained through bootstrapping with 1,000 samples, providing an assessment of variability.

The multimodal model accuracies for PorpoiseMMF ^17^ + Chowdhury signature ^9^ + CTransPath ^24^ setup is presented in Table. 1 as well as for the more granular TCGA genomics analysis in section 3. We investigated the MCAT ^20^ model + PRG (methods 3.6.3) + CTransPath ^24^ for deep exploration into WSI and proteomics interpretability (supplementary section 4.1-4.2).

### 3.3 Whole slide images

The PTRC-HGSOC dataset of 348 FFPE WSIs was downloaded from The Cancer Imaging Archive (TCIA) ^42^ [https://www.cancerimagingarchive.net/collection/ptrc-hgsoc/ representing 158 unique patients. The bookend FFPE slides were cut at 4 µm thickness using a microtome and mounted on glass slides (Leica Biosystems Cat 3800040) for H&E staining. Digital images of the H&E slides were recorded using a ScanScope AT Slide Scanner (Leica Aperio Technologies, Vista, CA, USA) under 20X objective magnification (0.4965 µm per pixel resolution). The 348 of the relevant WSIs of the TCGA-OV dataset that were part of the CPTAC-OV cohort were downloaded from The Cancer Imaging Archive (TCIA)^42^ https://www.cancerimagingarchive.net/collection/tcga-ov/ representing 175 unique patients TS1, TS2 (Fresh Frozen), DX1 and DX2 (FFPE) type slides. The TCGA-OV WSIs are heterogeneous in terms of scanner modalities, manufacturers, and acquisition protocols. In most cases, the images were acquired as part of routine care and not as part of a controlled research study or clinical trial. H&E slides were recorded under 20X objective magnification (0.5040 µm per pixel resolution).

### 3.4 Segmenting and embedding H&E WSIs

The images were visualized to ensure quality, segmented to remove the slide background or scanner artifacts with the CLAM ^21^ WSI preprocessing pipeline, and patched into 224 X 224 pixel patches representing approximately 112 X 112µm. To reduce the dimensionality of these large gigapixel images, these patches were then packed into tensor “bags” with dimensions (n,3,224,224) where n is the number of patches generated for a WSI. These patches were then embedded by the three feature-extracting vision transformer networks to create three separate sets of embeddings covering all samples and cohorts. The CTransPath ^24^ embedding model was chosen as was it best performing over a large set of tasks ^41^. The UNI^23^ model was chosen as it was pre-trained on non-TCGA data. This ensures there is no possible data leak when testing downstream models on TCGA WSI data.

Our DINO-OV model was also used as a third model to investigate if “same-domain” image embedders would provide a benefit for this task. Each model had an associated raw patch RGB space transformation and maps each patch dimensions (3, 224, 224) to a vector shaped (d) where d=1024 for UNI, d=384 for DINO-OV, and d=768 for CTransPath. The embedding stage was performed without stain normalization due to recent findings that this has little to no effect on downstream accuracy ^41^.

### 3.5 Proteomic Data

The TCGA MS proteomics data was downloaded from the Supplemental Information Table.3 of Zhang et al.^12^. Following pre-processing and validation from the original manuscript an average of 7,952 proteins were measured per tumor. The PTRC-HGSOC proteomics data was downloaded from the deposited data table: Processed LC-MRM-MS data ^9^. After pre-processing 8,800 proteins were found in over 50% of the measured bio-specimens.

The overlapping set of proteins from the TCGA and PTRC-HGSOC datasets were collected amounting to 7621 shared proteins. TCGA (CPTAC) and PTRC-HGSOC datasets were imputed with the scikit-learn KNNImputer ^43^, log normalized, and standard scaled to remove any batch effect present in the data. The alignment of datasets was validated by testing the overlap of distributions of each protein. We employed the nonparametric statistical test the Kolmogorov-Smirnov (KS) test, to compare the distribution of each protein between the two independent datasets. We found that they do not show a statistically significant difference: Mean KS Statistic: 0.112, Mean P-Value: 0.128.

### 3.6 Methodologies for proteomics groupings in multi-modal models

Deliberately selecting and structuring proteomics data to add to multi-modal deep learning models has an impact on final model performance as well as the inherent interpretability of the model once trained. We investigated proteomics selection with three high-level strategies. Signature-based, subtype-based, and pathways-based. A signatures-based strategy involves injecting only one fixed length 1d vector also known as a “proteomic signature” which is a subset of the proteomics data. This vector is found by feature selection methods and could be weights or unweighted. In our study, we investigated the unweighted “Chowdhury signature” (methods 3.6.1) proteomics signature. Subtypes-based grouping also offers another solution where non-overlapping protein “subtype” groups from structures (methods 3.6.2). Finally, pathways represent unique cellular functions, they constitute appropriate basic reasoning units suitable for interpretability. For multi-modal models, MCAT ^20^ and SurvPath ^19^data was organized into such groups (methods 3.6.3-3.6.4). We performed four different groupings to assess the importance and robustness of the multi-modal models to the groupings for this task. PorpoiseMMF models were trained with the same sets of proteins as these groups but not grouped instead as one vector input to be concatenated during model fusion. We experimented with two overlapping and two non-overlapping groupings to generate group-level tokens.

#### 3.6.1 Platinum-response protein signature (“Chowdhury Signature”)

Chowdhury et.al ^9^ describe a 64-gene unweighted proteomic signature that predicts response to platinum treatment. This signature was identified by investigation of 22 pathways based on 31 years of literature ^44^ followed by nonlinear machine-learning-based feature selection. From these proteins, 60 were measured in both our training and validation set. This signature was used as input to the PorpoiseMMF model. It was possible to group these proteins into 5 non-overlapping groups: Drug Metabolism & Biological Oxidation, Metabolic, Hypoxia, NK-kB as was described in Chowdhury et.al ^9^. These sub-groups were used in the MCAT and SurvPath architectures.

#### 3.6.2 CPTAC HGSOC subtypes (“CPTAC grouping”)

Zhang et.al ^12^ describe 1664 proteins clustered into 7 proteomic subtypes from the non-overlapping WGCNA-derived modules pathways based on the enrichment of KEGG and Reactome ontologies. These groups are labeled: ‘DNA replication’, ‘cell-cell communications’, ‘cytokine signaling’, ‘erythrocyte and platelet’, ‘ECM interaction’, ‘complement cascade’, and ‘metabolism’. This method aimed to inject subtype information into the model explicitly to see if it enhanced model accuracy.

#### 3.6.3 Platinum resistant genes and pathways (“PRG pathways”)

Huang et.al ^44^ curated a list of >900 proteins associated with platinum resistance in cancer from 30 years of literature. From these proteins, 587 were found for all patient samples in our studied cohorts. This is an appropriate list of pathways and genes to use when exploring HGSOC treatment responses as they cover a broad spectrum of biological processes implicated in drug resistance. Specifically, regulation of drug dynamics: the control over drug entry, exit, accumulation, sequestration, and detoxification within the cells, DNA damage response: enhanced capabilities for repairing and tolerating platinum-induced DNA damage, cell survival pathways: Alterations that allow cancer cells to avoid death despite chemotherapy, Pleiotropic processes: Changes in broad, multifunctional pathways that affect cancer cell behavior and finally, Tumor micro-environment: Variations in the cellular environment surrounding tumors that may contribute to drug resistance.

#### 3.6.4 Repair mechanism pathways (“RM pathways”)

To investigate the role of DNA repair mechanisms-related pathways in predicting response to platinum-treatment we utilized the 1664 DNA repair-related pathways from MSigDB and KEGG. These pathways are more mechanistic than the “PRG pathways” section 3.6.3 and are not constrained to the platinum response. See Figure. 5 for pathways and genes ranked by importance.

### 3.7 Models

#### 3.7.1 Training

For each experiment, we created a 5-fold split stratified on patient ID to ensure samples from the same patient were never found in the training and validation sets. Each learning task was set up as a binary classification (Sensitive vs Refractory). The proteomics-only baseline was an ensemble of ElasticNet, Random Forest, and XGBoost as described in Chowdhury et.al ^9^.

We trained all the deep learning based binary classification models for 15 epochs with gradient descent and the ADAM optimizer and a learning rate of 10^−4. CLAM and multimodal models were trained with a used a binary cross-entropy loss. Models were checkpointed during training and the checkpoints with the lowest validation loss were used for evaluation on test sets. We used L2 regularization with L = 10^−5 and weighted sampling to make sure models were not biased by class imbalances. We used a binary cross-entropy loss:

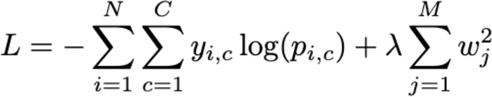

Where: N is the number of samples, C is the number of classes, yi. C is a binary indicator of whether class label c is the correct classification for observation i, pi. c is the predicted probability that observation i is of class c, λ is the regularization parameter, M is the number of weights, and wj represents each weight parameter in the model.

The full pipeline was trained on 3 Quadro RTX 8000 GPUS at the Budapest EMK server. The CLAM models were run with an SVM bag clustering loss. Metadata was checked for the PTRC-HGSOC hospital hold-out experiments to see if platinum sensitivity could be predicted from this data. Random forest models were not able to predict the label with AUC=0.47 for the UAB hold-out and AUC=0.55 for the Mayo hold-out experiment. This ruled out the learning of co-variate signals that could bias results or allow for shortcut learning.

## 4 Data availability

No new data was generated during this study. We release our splits of patients and cohorts in the supplementary information.

## 5 Code availability

The code to reproduce all results presented in the study is available at https://github.com/csabaiBio/HGSOC_platinum_response

## 6 Acknowledgements

This work was supported by the National Research, Development, and Innovation Office of Hungary within the framework of the MILAB Artificial Intelligence National Laboratory (RRF-2.3.1-21-2022-00004) and the Data-Driven Health Division of National Laboratory for Health Security (RRF-2.3.1-21-2022-00006) and by the European Union (Grant Agreement Nr. 10109571, SECURED Project) and OTKA K128780. Further support was provided by the Stipendium Hungaricum Program under the Tempus Public Foundation. It was also supported by the Research and Technology Innovation Fund (KTIA_NAP_13-2014-0021 and 2017-1.2.1-NKP-2017-00002 to Z.S.), Breast Cancer Research Foundation (BCRF-23-159 to Z.S.), Kræftens Bekæmpelse (R325-A18809 and R342-A19788 to Z.S.), Det Frie Forskningsråd Sundhed og Sygdom (2034-00205B to Z.S.), Basser Foundation (to Z.S.), NIH Grant 1 P01 CA228696-01A1 to Z.S.

## 7 Author contributions

Z.S, Zs.S, I.C and O.K conceived the experiments, O.K and Zs.S conducted the experiments. Z.S, Zs.S, I.C and O.K analyzed the results. A.O built out computational infrastructure for experiments. A.B trained the Ovarian-specific SSL WSI patch embedding model. P.P managed the computational infrastructure. L.M, Zs.S and analyzed the generated heatmaps. Z.S, Zs.S, and I.C supervised this project. All authors read and approved the final manuscript.

## 1. Supplementary Tables

**Table. S1.** Testing on TCGA samples AUC scores. Bold values are the highest scores for a given feature extractor and architecture. Underlined are the second-highest scores.

| HGSOC train TCGA test |  | Primary |  |  | Metastatic |  |  |
| --- | --- | --- | --- | --- | --- | --- | --- |
| Model |  | UNI | OV-Dino (ours) | CTransPath | UNI | OV-Dino | CTransPath |
| Multimodal | MCAT 60 | 0.69±0.047 | 0.675±0.049 | 0.668±0.049 | 0.618±0.051 | **0.667±0.049** | **0.704±0.047** |
|  | PorpoiseMMF 1k | 0.652±0.053 | 0.659±0.053 | 0.658±0.057 | **0.665±0.051** | 0.639±0.047 | <u>0.692±0.051</u> |
|  | PorpoiseMMF 60 | **0.751±0.045** | **0.742±0.046** | **0.721±0.046** | <u>0.656±0.05</u> | <u>0.641±0.052</u> | 0.0±0.0 |
|  | SurvPath 60 | <u>0.708±0.047</u> | <u>0.688±0.049</u> | <u>0.69±0.049</u> | 0.627±0.048 | 0.629±0.053 | 0.619±0.054 |
| Omics | 1k protein ensemble | 0.536±0.054 | 0.536±0.054 | 0.536±0.054 | 0.599±0.056 | 0.599±0.056 | 0.599±0.056 |
|  | 60 protein ensemble | 0.61±0.05 | 0.61±0.05 | 0.61±0.05 | 0.54±0.053 | 0.54±0.053 | 0.54±0.053 |
| WSI | clam_sb | 0.544±0.053 | 0.566±0.051 | 0.542±0.054 | 0.507±0.055 | 0.45±0.052 | 0.486±0.054 |

**Table. S2.**
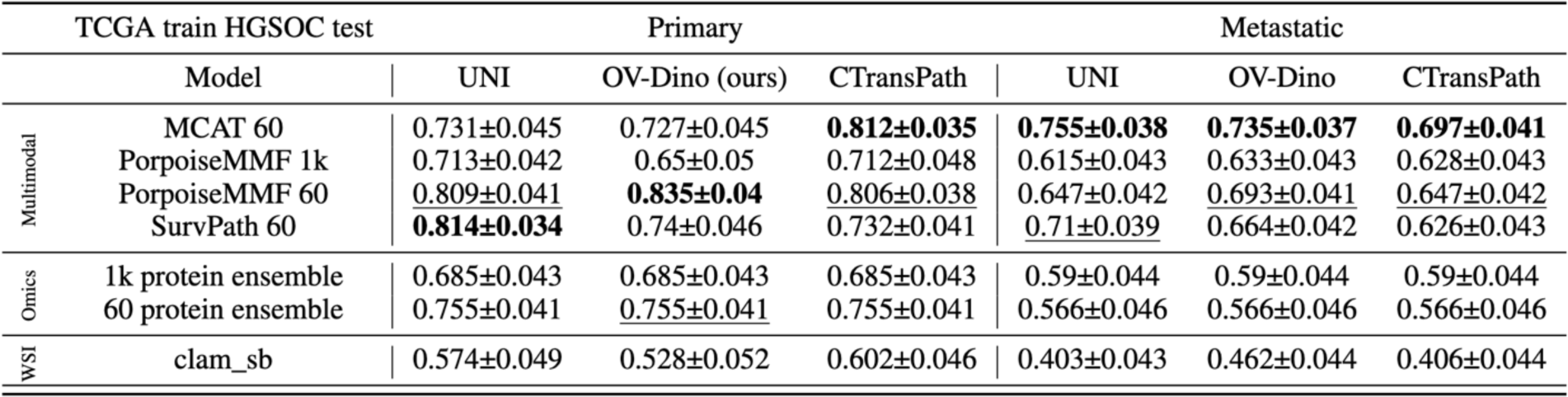
Testing on all HGSOC samples AUC scores. Bold values are the highest scores for a given feature extractor and architecture. Underlined are the second-highest scores.

| TCGA train HGSOC test |  | Primary |  |  | Metastatic |  |  |
| --- | --- | --- | --- | --- | --- | --- | --- |
| Model |  | UNI | OV-Dino (ours) | CTransPath | UNI | OV-Dino | CTransPath |
| Multimodal | MCAT 60 | 0.731±0.045 | 0.727±0.045 | **0.812±0.035** | **0.755±0.038** | **0.735±0.037** | **0.697±0.041** |
|  | PorpoiseMMF 1k | 0.713±0.042 | 0.65±0.05 | 0.712±0.048 | 0.615±0.043 | 0.633±0.043 | 0.628±0.043 |
|  | PorpoiseMMF 60 | <u>0.809±0.041</u> | **0.835±0.04** | <u>0.806±0.038</u> | 0.647±0.042 | <u>0.693±0.041</u> | <u>0.647±0.042</u> |
|  | SurvPath 60 | **0.814±0.034** | 0.74±0.046 | 0.732±0.041 | <u>0.71±0.039</u> | 0.664±0.042 | 0.626±0.043 |
| Omics | 1k protein ensemble | 0.685±0.043 | 0.685±0.043 | 0.685±0.043 | 0.59±0.044 | 0.59±0.044 | 0.59±0.044 |
|  | 60 protein ensemble | 0.755±0.041 | <u>0.755±0.041</u> | 0.755±0.041 | 0.566±0.046 | 0.566±0.046 | 0.566±0.046 |
| WSI | clam_sb | 0.574±0.049 | 0.528±0.052 | 0.602±0.046 | 0.403±0.043 | 0.462±0.044 | 0.406±0.044 |

**Table. S3.** Testing on HGSOC-UAB samples AUC scores. Bold values are the highest scores for a given feature extractor and architecture. Underlined are the second-highest scores.

| HGSOC UAB hold out |  | Primary |  |  | Metastatic |  |  |
| --- | --- | --- | --- | --- | --- | --- | --- |
| Model |  | UNI | OV-Dino (ours) | CTransPath | UNI | OV-Dino | CTransPath |
| Multimodal | MCAT 60 | 0.688±0.092 | 0.693±0.104 | 0.802±0.08 | 0.639±0.085 | **0.765±0.077** | <u>0.798±0.065</u> |
|  | PorpoiseMMF 1k | 0.615±0.097 | 0.727±0.085 | 0.441±0.108 | 0.647±0.081 | 0.622±0.083 | 0.706±0.078 |
|  | PorpoiseMMF 60 | <u>0.823±0.072</u> | **0.84±0.07** | **0.88±0.069** | 0.567±0.089 | 0.662±0.081 | 0.649±0.092 |
|  | SurvPath 60 | **0.826±0.069** | <u>0.729±0.08</u> | <u>0.852±0.058</u> | **0.729±0.071** | <u>0.676±0.078</u> | **0.832±0.06** |
| Omics | 1k protein ensemble | 0.476±0.103 | 0.476±0.103 | 0.476±0.103 | 0.355±0.08 | 0.355±0.08 | 0.355±0.08 |
|  | 60 protein ensemble | 0.558±0.094 | 0.558±0.094 | 0.558±0.094 | <u>0.665±0.092</u> | 0.665±0.092 | 0.665±0.092 |
| WSI | clam_sb | 0.406±0.11 | 0.451±0.107 | 0.539±0.088 | 0.546±0.087 | 0.538±0.091 | 0.769±0.081 |

**Table. S4.** Testing on HGSOC-Mayo samples AUC scores. Bold values are the highest scores for a given feature extractor and architecture. Underlined are the second-highest score.

| HGSOC MAYO hold out |  | Primary |  |  | Metastatic |  |  |
| --- | --- | --- | --- | --- | --- | --- | --- |
| Model |  | UNI | OV-Dino (ours) | CTransPath | UNI | OV-Dino | CTransPath |
| Multimodal | MCAT 60 | 0.61±0.095 | 0.675±0.09 | 0.61±0.106 | 0.79±0.064 | 0.75±0.062 | 0.758±0.062 |
|  | PorpoiseMMF 1k | 0.523±0.105 | 0.455±0.104 | 0.422±0.097 | 0.812±0.052 | <u>0.849±0.048</u> | 0.817±0.052 |
|  | PorpoiseMMF 60 | **0.802±0.075** | <u>0.75±0.091</u> | <u>0.698±0.09</u> | **0.877±0.042** | **0.898±0.037** | **0.872±0.042** |
|  | SurvPath 60 | <u>0.659±0.098</u> | 0.662±0.094 | **0.731±0.087** | 0.812±0.054 | 0.759±0.062 | 0.814±0.057 |
| Omics | 1k protein ensemble | 0.4±0.094 | 0.4±0.094 | 0.4±0.094 | <u>0.83±0.057</u> | 0.83±0.057 | <u>0.83±0.057</u> |
|  | 60 protein ensemble | 0.539±0.1 | 0.539±0.1 | 0.539±0.1 | 0.792±0.057 | 0.792±0.057 | 0.792±0.057 |
| WSI | clam_sb | 0.545±0.098 | **0.766±0.078** | 0.481±0.107 | 0.653±0.07 | 0.526±0.075 | 0.459±0.074 |

**Table S5.** Opensource datasets used for OV-Dino ViT pre-training.

| **Dataset** | **Number of WSIs** | **Micron per pixel** | **slide type** |
| --- | --- | --- | --- |
| TCGA-OV <sup>42</sup> | 1481 | 0.5040 | FFPE + Fresh frozen |
| Bevacizumab response<sup>45</sup> | 284 | 0.5 | FFPE |
| CPTAC 2 <sup>12</sup> | 221 | 0.2501 | FFPE |
| UBC Ocean <sup>42</sup> | 538 | 0.5 | FFPE |
| PTRC-HGSOC <sup>9</sup> | 349 | 0.4965 | FFPE |

## 2. Supplementary Figures

**Figure. S1.**
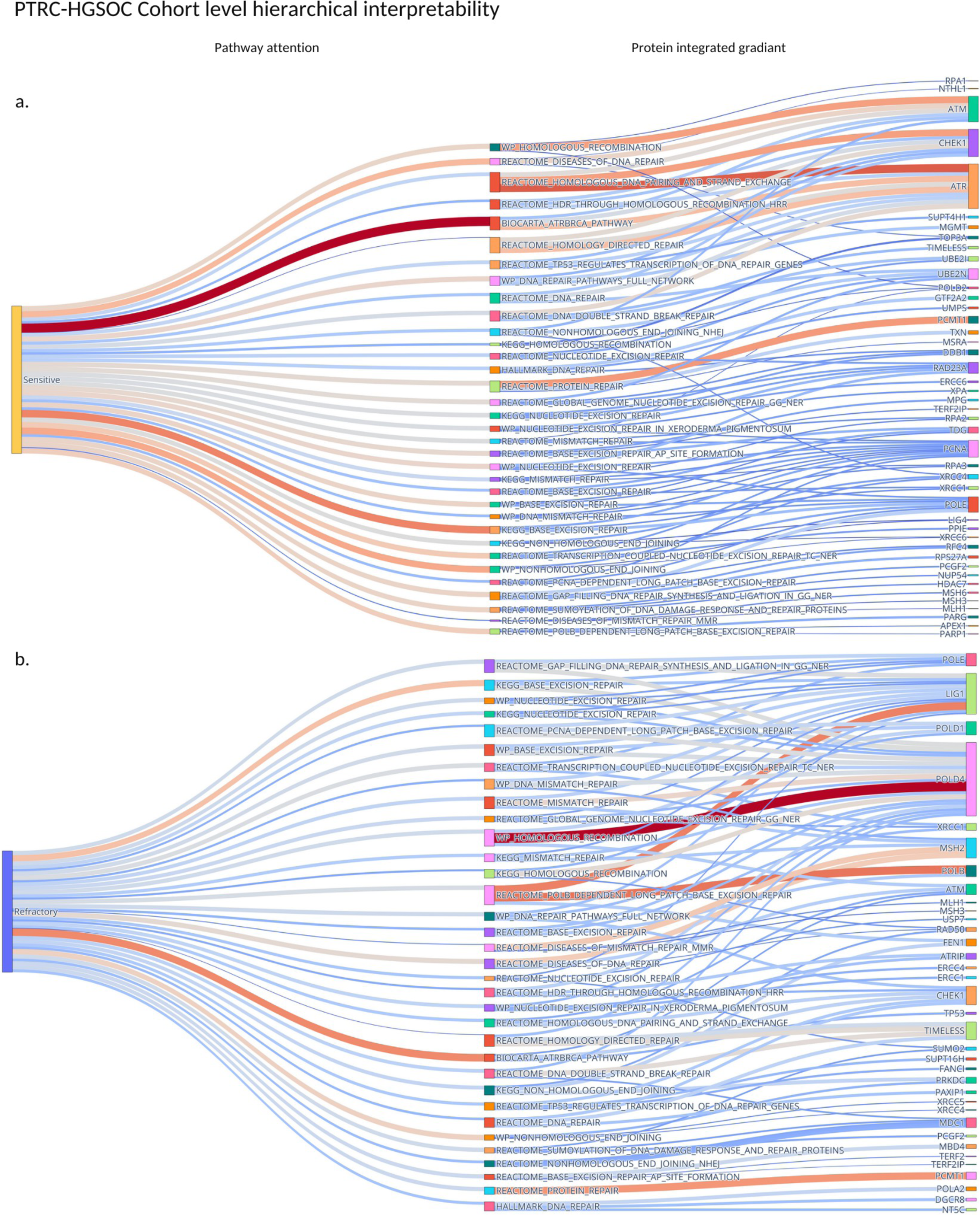
Global interpretability for entire cohorts for the MCAT model. Sankey diagrams where weights connecting cohorts to pathways are the mean model weights. Connections between pathways and proteins represent the four proteins in each given pathway with the largest integrated gradients. Wider connecting lines are higher integrated gradients important. This offers a hierarchic interpretability. **a.** Sensitive pathways and proteins with high attention, **b.** Refractory pathways and proteins with high attention.

**Figure. S2.**
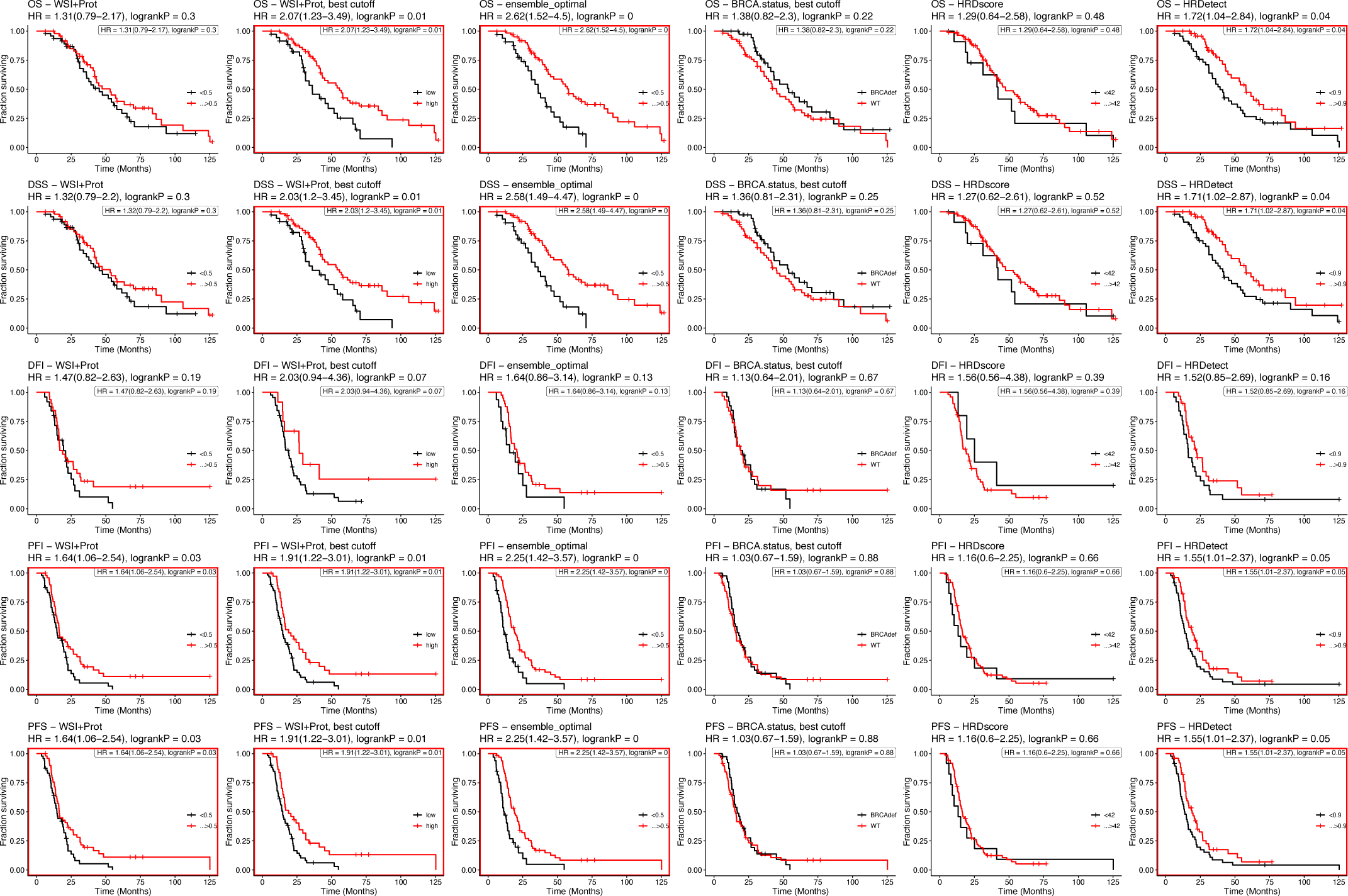
Survival analysis for OS, DSS, DFI, PFI, PFS for TCGA-OV as a test cohort with genetic data available.

**Figure. S3.**
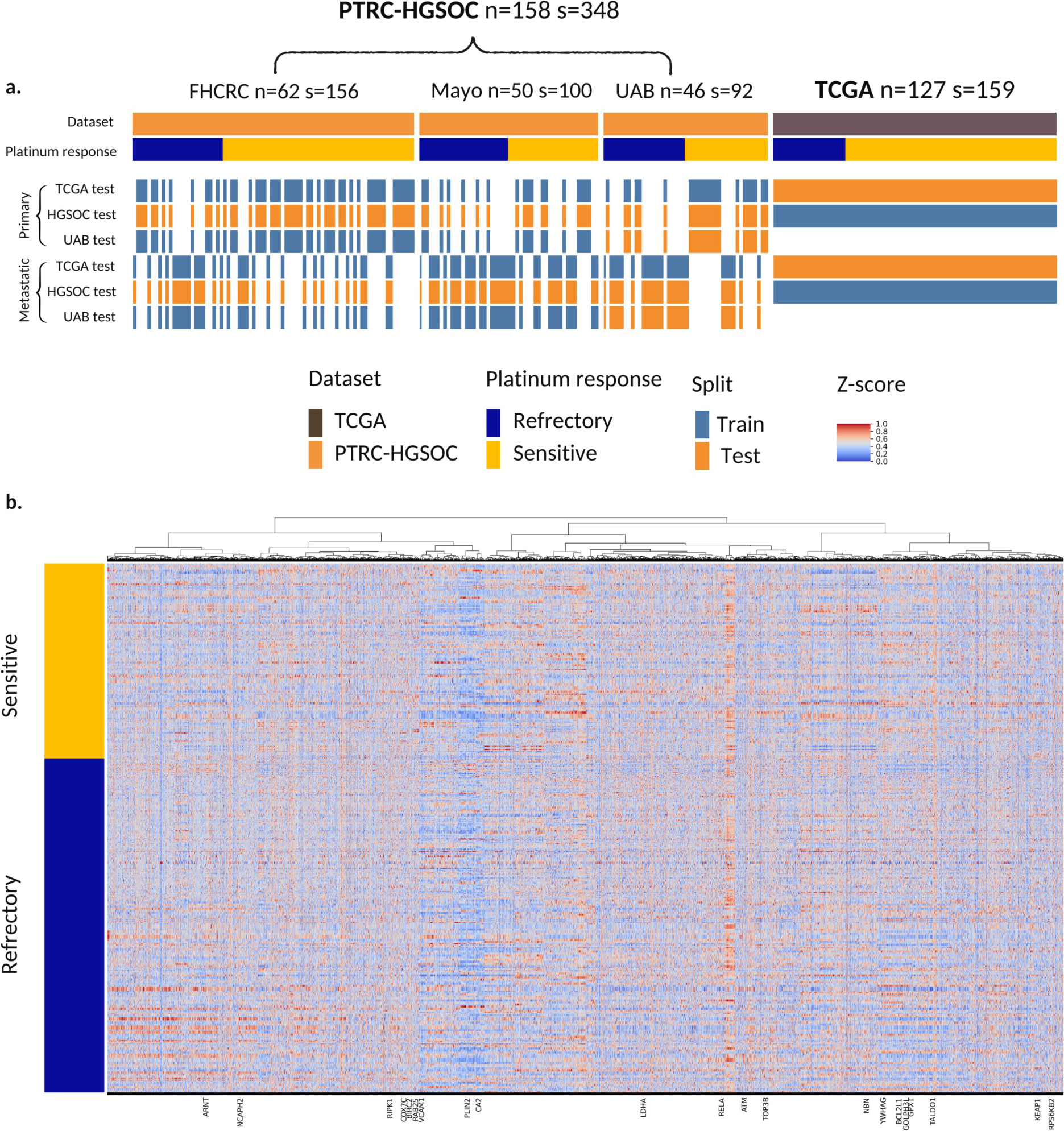
Data overview for the study. **a.** cohorts and splits used for experimental validation. **b.** Hierarchical clustering of 7567 proteomics measurements available for each patient

**Figure S4.**
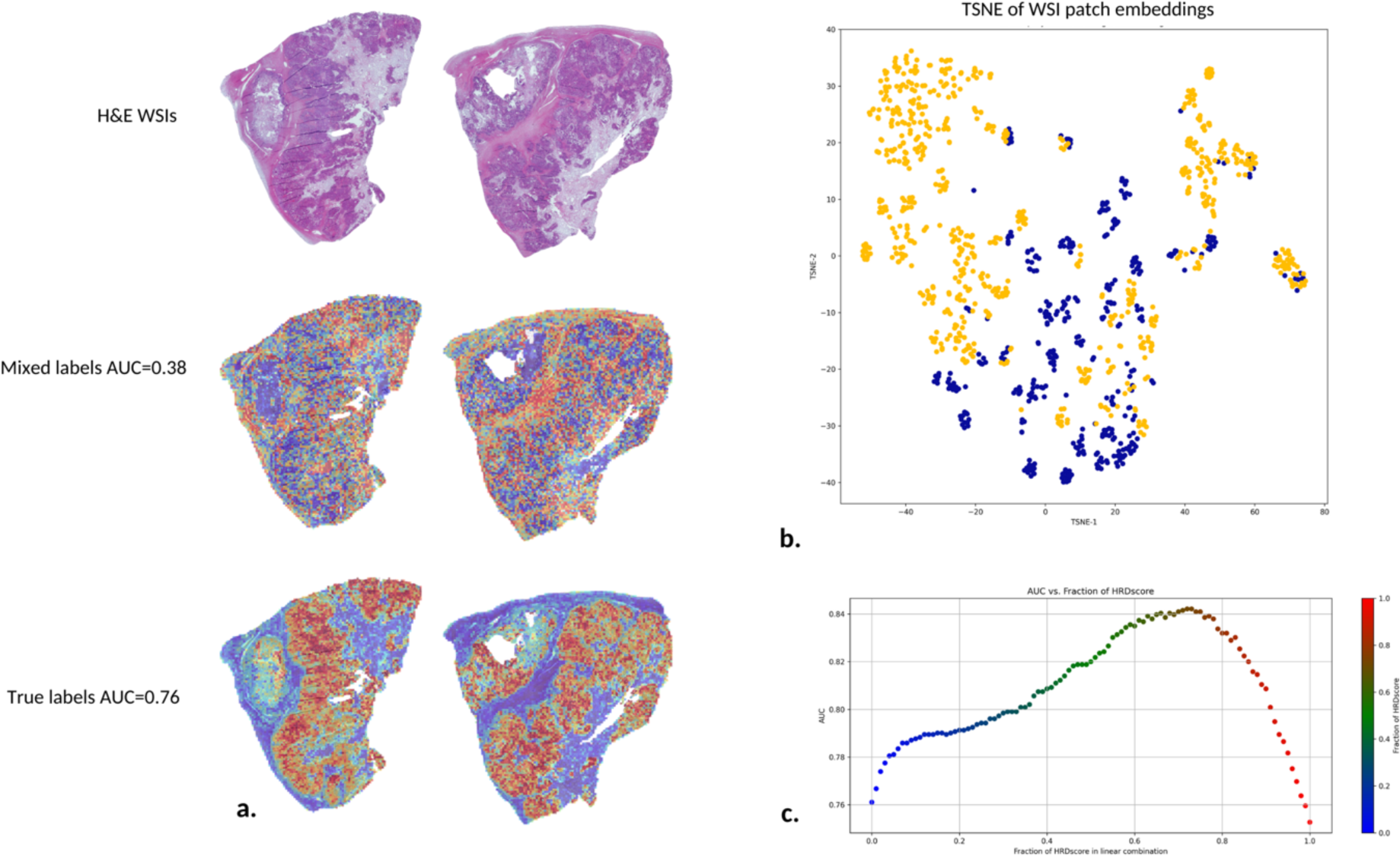
**a.** We retrained the MCAT model (same as in methods 3.7) with deliberately randomly mixed labels to assess how heatmaps would degrade with model performance. Heatmaps are dispersed in models trained with the deliberately mixed and therefore incorrect labels. They highlight meaningful areas of the WSI for trained models. **b.** TSNE of UNI top 1 embeddings for each sensitive and refractory groups clustering indicated the model found distinct morphologies for sensitive (yellow points) and refractory (blue points) patients. **c**. AUC variation with varying mixing of HRDscore and our WSI + proteomics model to create the “Ensemble” linear combination model (Figure 3a, Figure 3.b).

## 3. Supplementary methods

### 3.1 Multiple instance learning

Multiple instance learning (MIL) is a popular framework based on modeling set-based structures called “bags”. Each bag has a label and “instances” inside this bag do not. A bag label is defined by the unknown labels of its containing instances. One or more instances in a bag define its label. The goal of classifying a bag from its instances is also known as “weakly supervised learning”. MIL is a natural modeling choice for WSIs which are gigapixel-sized images. Patching up the images into “instances” xi with dimensions (3×224×224) allows for the generation of bags with dimensions (n×3×224×224) where n is the number of patches in the whole slide image and 3 represents the RGB channels of the light microscopy high-resolution image. The objective of MIL is to learn a function that maps bags to labels with a model composed of instance-level functions that are aggregated with a symmetric, permutation-invariant aggregation function that pools the extracted features into a single bag-level feature embedding. This embedding can then be mapped to the WSI “bag” label ^21^.

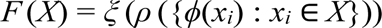

Where F is the function to learn, X is the bag data. ρ is the aggregation function and ϕ is the instance level function. ξ is a bag-level classifier that maps the whole WSI embedding to the clinical endpoint also known as the label in the learning task (sensitive or refractory in our case).

### 3.2 Multi-modal deep learning

The integration of multi-modal data such as Whole Slide Images (WSIs) and proteomics is achieved through the modeling of the joint probability distribution, P (WSIs, Proteomics). This framework underpins the ability to harness complementary and independent information from each modality, enhancing the predictive power of models. Specifically, by constructing a function f that leverages this joint distribution to map input data to a target variable Y, in our case treatment response (Y = f (WSIs, Proteomics)), we exploit the comprehensive insights provided by each data type. The modeling process involves probabilistic inference to estimate the posterior distribution P (Y WSIs, Proteomics), applying Bayes’ theorem to integrate prior knowledge with empirical data. This approach not only facilitates the extraction of nuanced, modality-specific features but also illuminates the complex interdependencies between different data types, advancing the precision and cross-modality interpretability of predictions in personalized medicine. The multimodal architectures we have used were built for WSI + RNAseq data but we have adapted them for WSI + proteomics.

### 3.3 Clustering-constrained Attention Multiple Instance Learning (CLAM)

CLAM^21^ is a framework to perform WSI-only MIL (supplementary 3.1) with an attention-based pooling function in the place of *ρ*.

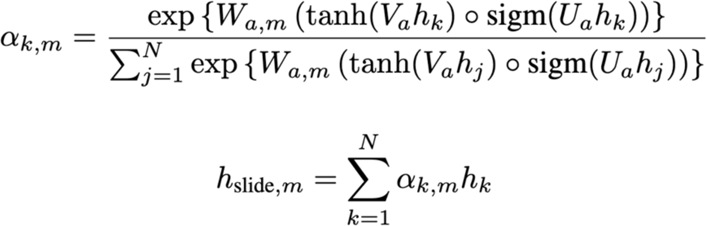

W1 is the first fully-connected layer that compresses each 1024-dimensional patch-level representation into a 512-dimensional vector. Ua is the first layer of the attention network, part of the attention backbone shared by all classes. Va is the second layer of the attention network, part of the attention backbone shared by all classes. Wa,1 to Wa,n are the n parallel attention branches in the attention network. Wc,1 to Wc,n are the n parallel, independent classifiers used to score each class-specific slide-level representation. *α* k,m is the attention score of the k-th patch for the m-th class. This model is also “fortified” with an Instance-level Clustering mechanism. This is designed to further encourage the learning of class-specific features. We used the SVM loss in our CLAM baseline. Full implementation can be found at https://github.com/mahmoodlab/CLAM.

### 3.4 Pathology-Omics Research Platform for Integrative Survival Estimation (PorpoiseMMF)

PorrpoiseMMF^18,19^ is a late fusion model that combines MIL and an omics vector channel to preform multimodal deep learning. WSI and Omics channels are encoded to fixed size internal representations. Then the Kronecker Product between WSI embeddings and proteomic embeddings is taken before the combined representation is passed through further layers. Full implementation can be found at https://github.com/mahmoodlab/PORPOISE.

### 3.5 Multi-modal Co-Attention Transformer (MCAT)

MCAT^20^ is an early fusion model where patch embeddings and pathway embeddings are fused with a co-attention mechanism. This allows for sharing of information between various parts of the input early in the network. Unlike PorpoiseMMF, WSI patches and pathways may more directly interact and modulate one another. This is achieved by a cross-attention module to model the attention of histology patches (keys K, values V) toward gene sets (queries Q):

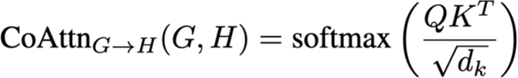

Full implementation can be found at https://github.com/mahmoodlab/MCAT.

### 3.6 Biological Pathways and Histology for Survival Prediction (SurvPath)

SurvPath^19^ is an early fusion model Early that can model dense multi-modal interactions between pathway and patch tokens. This model aims to build on MCAT. SurvPath allows for genes to patch interactions as well and patch to genes interactions as well as a more fine-grained tokenization process. This model employs a transformer attention ^19^ that can measure and aggregate pair-wise interactions between multi-modal tokens:

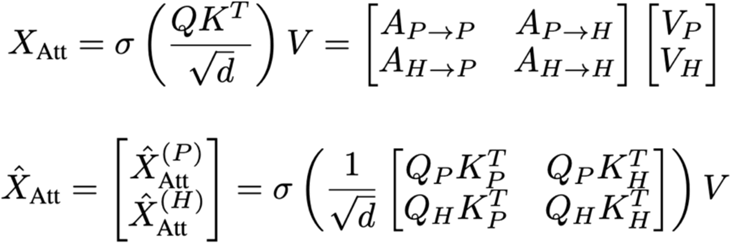

Where A values are self-attention, QP and QH denote the subset of pathway and histology queries and keys.

Full implementation can be found at https://github.com/mahmoodlab/SurvPath.

### 3.7 Integrated gradients for protein level importance

We used the gradient-based feature attribution method of Integrated gradients ^28^ to understand which features (protein expression values) have the largest impact on the model output (tumor response). Larger values push to model to have larger outputs (more sensitive). We define **x**^′^ as the baseline input (zeros in the shape of the model input in our case) and *x* as the feature, *F* is the trained model. A “straight line path” from **x**^′^ to *x* is generated in steps *α* and we calculated all the gradients of *F* along this path. The Integrated gradients are defined as the path integral over all of these gradients accumulating them to generate an interpretable, sensitive, and implementation invariant importance value for each input. This method allows for further model interpretability and understanding of the learned importance of proteins as well as the extent to which they are implicated as relevant to the mechanism or pathway of tumor sensitivity.

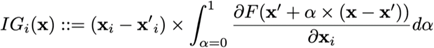

### 3.8 Feature extractors

#### 3.8.1 SSL DINO-OV

Intending to optimally extract Ovarian WSI patch features for downstream learning, we trained a patch embedding network based on 5 open-source Ovarian cancer H&E WSI datasets with Y total images at maximum magnification “level 0” (20X magnification) to create a diverse dataset of 16.2 million images. These were used to train a ViT neural network with self-distillation with no labels (DINO) ^46^ in a self-supervised manner. DINO is an algorithm for visual representation working to distill knowledge from a teacher network to a student network by minimizing the Kullback-Leibler (KL) divergence between their predicted probability distributions for various augmentations (global and local) of the same input image.

## 4. Supplementary discussion

### 4.1 Co-attention heatmaps

In a typical image recognition scenario, rather than being directly provided with a bounding box indicating the location of the animal, the machine learning model is given a collection of photographs with captions to learn to recognize different animals. Each photo might show an animal in various settings, and the caption briefly describes what’s happening in the image. During the training, from correlations between image features and captions, the model notices patterns. Every time the word “cat” appears in the caption, the photo features a furry creature with pointed ears and a tail. Over time, by associating words in the captions with visual features (textures, edges, corners, etc.) in the images, the model learns to identify cats in any photo, regardless of the setting or pose, without needing explicit annotations for “cat” within the image ^47^.

In our study, we apply a similar approach to understand histopathology whole slide images of tissues stained with H&E, paired with corresponding proteomics data. Think of the histopathology images as the photographs and the proteomics data as the captions. The proteomics data describe the molecular “story” of the tissue, such as which proteins are present and in what abundance, indicating which molecular pathways are active. By training a multimodal AI network on these paired datasets, the network learns to associate specific visual patterns in the histopathology images (like the arrangement and characteristics of cells and tissues) with the active molecular pathways described by the proteomics data. Over time, just as a simple model learned to identify cats by their visual features from photos with captions, the AI learns to “see” which molecular pathways are active in different parts of the tissue by analyzing its visual features. The attention maps generated by the AI model are akin to highlighting the “cats” in our analogy; they color-code the tissue images to indicate where and which pathways are active, allowing us to visually interpret complex molecular activities in the context of tissue structure and morphology.

### 4.2 Extended discussion: Early vs late fusion?

To decide between early vs late fusion as the optimal choice, late fusion with a pre-defined proteomic signature seems to perform best, however early fusion models still perform very well and have the added benefit of a fully comprehensive interpretability analysis. In terms of how the input structured and model architectures can be chosen, these modeling choices are all interrelated and have complex effects on one another. For example, the choice of protein input and patch featurization model affects the number of model parameters. These covariances make it difficult to conclude the best choice of architecture, protein input, and embedding beyond pure empirical study. However, our results indicate that a proteomics machine-learning-based feature selection followed by the simplest late fusion concatenation model PorpoiseMMF led to the overall most robust performance. This may be as late fusion facilitates a more tailored feature extraction and representation learning for each separate modality. This may ensure that the model can leverage the strengths of each data source better handling disparities in the dimensionality, scale, and quality across modalities, leading to a more robust and accurate synthesis of information for decision-making. This finding is contrary to the publications of the methods ^19^. However, early fusion may outperform late fusion in the larger data regime. This is where the models were benchmarked at over 1000 samples per class, almost an order of magnitude larger scale than the available HGSOC response data we studied. Despite having lower AUC scores in our study, early fusion MCAT and SurvPath models as they allow for the in-depth interpretability analysis we present. Such interpretability advantages may help comply with GDPR-related tests that need to take place before clinical deployment.

